# Efficacy of Colchicine in Non-Hospitalized Patients with COVID-19

**DOI:** 10.1101/2021.01.26.21250494

**Authors:** Jean-Claude Tardif, Nadia Bouabdallaoui, Philippe L. L’Allier, Daniel Gaudet, Binita Shah, Michael H. Pillinger, Jose Lopez-Sendon, Protasio da Luz, Lucie Verret, Sylvia Audet, Jocelyn Dupuis, André Denault, Martin Pelletier, Philippe A. Tessier, Sarah Samson, Denis Fortin, Jean-Daniel Tardif, David Busseuil, Elisabeth Goulet, Chantal Lacoste, Anick Dubois, Avni Y. Joshi, David D. Waters, Priscilla Hsue, Norman E. Lepor, Frédéric Lesage, Nicolas Sainturet, Eve Roy-Clavel, Zohar Bassevitch, Andreas Orfanos, Jean C. Grégoire, Lambert Busque, Christian Lavallée, Pierre-Olivier Hétu, Jean-Sébastien Paquette, Sylvie Levesque, Mariève Cossette, Anna Nozza, Malorie Chabot-Blanchet, Marie-Pierre Dubé, Marie-Claude Guertin, Guy Boivin, for the COLCORONA Investigators

## Abstract

**Background:** Evidence suggests the role of an inflammatory storm in COVID-19 complications. Colchicine is an orally administered, anti-inflammatory medication beneficial in gout, pericarditis and coronary disease.

**Methods:** We performed a randomized, double-blind trial involving non-hospitalized patients with COVID-19 diagnosed by polymerase chain reaction (PCR) testing or clinical criteria. The patients were randomly assigned to receive colchicine (0.5 mg twice daily for 3 days and once daily thereafter) or placebo for 30 days. The primary efficacy endpoint was the composite of death or hospitalization for COVID-19.

**Results:** A total of 4488 patients were enrolled. The primary endpoint occurred in 4.7% of the patients in the colchicine group and 5.8% of those in the placebo group (odds ratio, 0.79; 95.1% confidence interval (CI), 0.61 to 1.03; P=0.08). Among the 4159 patients with PCR-confirmed COVID-19, the primary endpoint occurred in 4.6% and 6.0% of patients in the colchicine and placebo groups, respectively (odds ratio, 0.75; 95% CI, 0.57 to 0.99; P=0.04). In these patients with PCR-confirmed COVID-19, the odds ratios were 0.75 (95% CI, 0.57 to 0.99) for hospitalization due to COVID-19, 0.50 (95% CI, 0.23 to 1.07) for mechanical ventilation, and 0.56 (95% CI, 0.19 to 1.66) for death. Serious adverse events were reported in 4.9% and 6.3% in the colchicine and placebo groups (P=0.05); pneumonia occurred in 2.9% and 4.1% of patients (P=0.02). Diarrhea was reported in 13.7% and 7.3% in the colchicine and placebo groups (P<0.0001).

**Conclusion:** Among non-hospitalized patients with COVID-19, colchicine reduces the composite rate of death or hospitalization. (COLCORONA ClinicalTrials.gov number: NCT04322682)

---

Accumulating evidence suggests that some patients with coronavirus disease 2019 (COVID-19) suffer from the cytokine storm syndrome.^1^ Treatment of this exaggerated inflammatory response has been advocated to address the immediate need to reduce the risk of complications.^1,2^ The steroid dexamethasone reduces mortality in patients hospitalized with COVID-19, but only if they receive mechanical ventilation or supplemental oxygen.^3^ In addition, the anti-interleukin-6 receptor antibody tocilizumab was shown to reduce the likelihood of progression to mechanical ventilation in patients hospitalized for COVID-19 pneumonia.^4^

The severe acute respiratory syndrome coronavirus 1 (SARS-CoV-1), which is closely related to the SARS-CoV-2 virus responsible for COVID-19, has been shown to activate the NLRP3 inflammasome.^5^ This intracellular complex activates several interleukins, which then trigger an inflammatory cascade. Given that elevated levels of interleukin-6 are associated with adverse clinical outcomes in COVID-19,^6^ targeting the NLRP3 inflammasome may reduce related complications in patients at risk of cytokine activation.

Prevention of COVID-19 complications in an out-patient setting ideally requires a clinically available, orally administered and inexpensive medication targeting the inflammasome with a known favorable safety and tolerability profile. Colchicine is a potent anti-inflammatory agent used to treat gout, viral pericarditis, coronary disease and familial Mediterranean fever.^7-10^ Its mechanism of action is through the inhibition of tubulin polymerization, with effects on the inflammasome, cellular adhesion molecules and inflammatory chemokines.^11-13^ In an experimental model of acute respiratory distress syndrome, colchicine was shown to reduce inflammatory lung injury and respiratory failure by interfering with leukocyte activation and recruitment.^14^

We conducted the COLchicine CORONAvirus SARS-CoV-2 (COLCORONA) trial in non-hospitalized patients with COVID-19 to evaluate the effects of colchicine on complications including hospitalization and death and its safety and tolerability

## METHODS

### TRIAL DESIGN

COLCORONA was a randomized, double-blind, placebo-controlled, investigator-initiated trial comparing colchicine (0.5 mg twice daily for the first 3 days and then once daily for 27 days thereafter) with placebo in a 1:1 ratio. The study was funded by the Government of Quebec, the Bill and Melinda Gates Foundation, the National Institutes of Health and philanthropist Sophie Desmarais. The trial protocol, available with the full text of this article at XX.org, was designed by the study steering committee. The protocol was approved by the institutional review board at all centers involved in the 6 countries that participated in the trial (Supplementary Appendix). All study support activities, including project coordination, data management, site monitoring, and statistical oversight and analyses, were performed at the Montreal Health Innovations Coordinating Center (MHICC). The trial was overseen by a data safety monitoring board of independent experts. The study medication and matching placebo were provided by Pharmascience (Montreal), which had no role in the design or conduct of the trial or the preparation or review of the manuscript. The first author (JCT) and lead statistician (MCG) prepared the first draft of the manuscript, had full access to the trial database, generated statistical analyses, made the decision to submit the manuscript for publication, and assume responsibility for the accuracy and completeness of the data and analyses and for the fidelity to the protocol.

### TRIAL POPULATION

Patients were eligible if they were at least 40 years of age, had received a diagnosis of COVID-19 within 24 hours of enrollment, were not currently hospitalized and not under immediate consideration for hospitalization, and presented at least one of the following high-risk criteria: age of 70 years or older, obesity (body-mass index of 30 kg/m^2^ or more), diabetes, uncontrolled hypertension (systolic blood pressure ≥150 mm Hg), known respiratory disease, known heart failure, known coronary disease, fever of at least 38.4°C within the last 48 hours, dyspnea at the time of presentation, bicytopenia, pancytopenia, or the combination of high neutrophil and low lymphocyte counts. The diagnosis of COVID-19 was made by local laboratories using polymerase chain reaction testing on a naso-pharyngeal swab specimen. Given the restrictions in laboratory testing early in the pandemic, a diagnosis was also accepted as an epidemiological link with a household member who had a positive nasopharyngeal test result for patients with symptoms compatible with COVID-19, or by a clinical algorithm in a symptomatic patient without an obvious alternative cause as per official guidelines (Supplementary Table S1).^15^ Women were either not of childbearing potential or practicing adequate contraception.

Patients were excluded if they had inflammatory bowel disease, chronic diarrhea or malabsorption; pre-existent progressive neuromuscular disease; estimated glomerular filtration rate less than 30 ml/minute/1.73 m^2^; severe liver disease; current treatment with colchicine; current chemotherapy for cancer; or a history of significant sensitivity to colchicine. Further details regarding eligibility criteria are provided in the Supplementary Appendix.

Written informed consent was obtained electronically or on paper from all patients before enrollment following a telemedicine or in-person visit, respectively. Study medication was delivered at the patient’s house within 4 hours of enrollment. Clinical evaluations occurred by telephone at 15 and 30 days following randomization.

### ENDPOINTS

The primary efficacy endpoint was a composite of death or hospitalization due to COVID-19 infection in the 30 days following randomization. The secondary endpoints consisted of the components of the composite primary endpoint; and the need for mechanical ventilation in the 30 days following randomization. Pneumonias, other serious adverse events, and non-serious adverse events were also collected.

### STATISTICAL ANALYSES

It was estimated that a sample size of approximately 6000 randomized patients with 3000 patients in each treatment group would be required to detect a 25% relative risk reduction with colchicine with a power of 80% given a primary endpoint event rate of 7% in the placebo group and a two-sided test at the 0.05 significance level.

The efficacy analyses were conducted according to the intention-to-treat principle. The primary endpoint was compared between the two treatment groups using a chi-square test and the odds ratio along with 95.1% confidence interval, was provided. Secondary endpoints were analyzed similarly. Because of potential limitations to the specificity of COVID-19 diagnosis made on clinical or epidemiological criteria alone, a pre-specified analysis included only those patients who were enrolled based on a positive polymerase chain reaction test. Pre-specified subgroup analyses were conducted using logistic regression models including the treatment group, the subgroup factor and the treatment x subgroup factor interaction.

Interim analyses were planned after 25, 50 and 75% of the primary endpoint events had occurred. The pre-specified stopping rule for efficacy was based on the Lan-DeMets procedure with the O’Brien-Fleming alpha-spending function. Following its review of the first two interim results, the monitoring board recommended that the trial should continue as planned. On December 11, 2020, the steering committee chairman informed the data safety monitoring board that the investigators had decided to terminate the study once 75% of the planned patients were recruited and had completed the 30-day follow-up. This decision was made due to logistical issues related to maintaining the central study call center active 24 hours per day for a prolonged period of time, as well as the need to provide healthcare systems with study results in a timely fashion given the state of the COVID-19 pandemic. To account for the interim analyses, the statistical significance level was set to 0.0490 for the final analysis of the primary endpoint. All other statistical tests were two-sided and conducted at the 0.05 significance level.

Statistical analyses were performed using SAS version 9.4. There was no prespecified plan to adjust for multiple comparisons across the multiple methods used to analyze the primary outcome and secondary endpoints; results of these analyses are reported with point estimates and 95% CI, without P-values. 95% CIs are not adjusted for multiple comparisons and inferences drawn from them may not be reproducible. The statistical analysis plan was approved on November 25, 2020.

## RESULTS

### PATIENTS

Trial enrollment began in March 2020 and was completed in December 2020; the last trial visit was in January 2021. A total of 4488 patients underwent randomization and were followed for 30 days. At the time of database lock and unblinding on January 20, 2021, vital and primary endpoint event status were available for all except for 93 patients (97.9%). Details regarding the disposition of the patients are provided in Figure 1.

**Figure 1.**
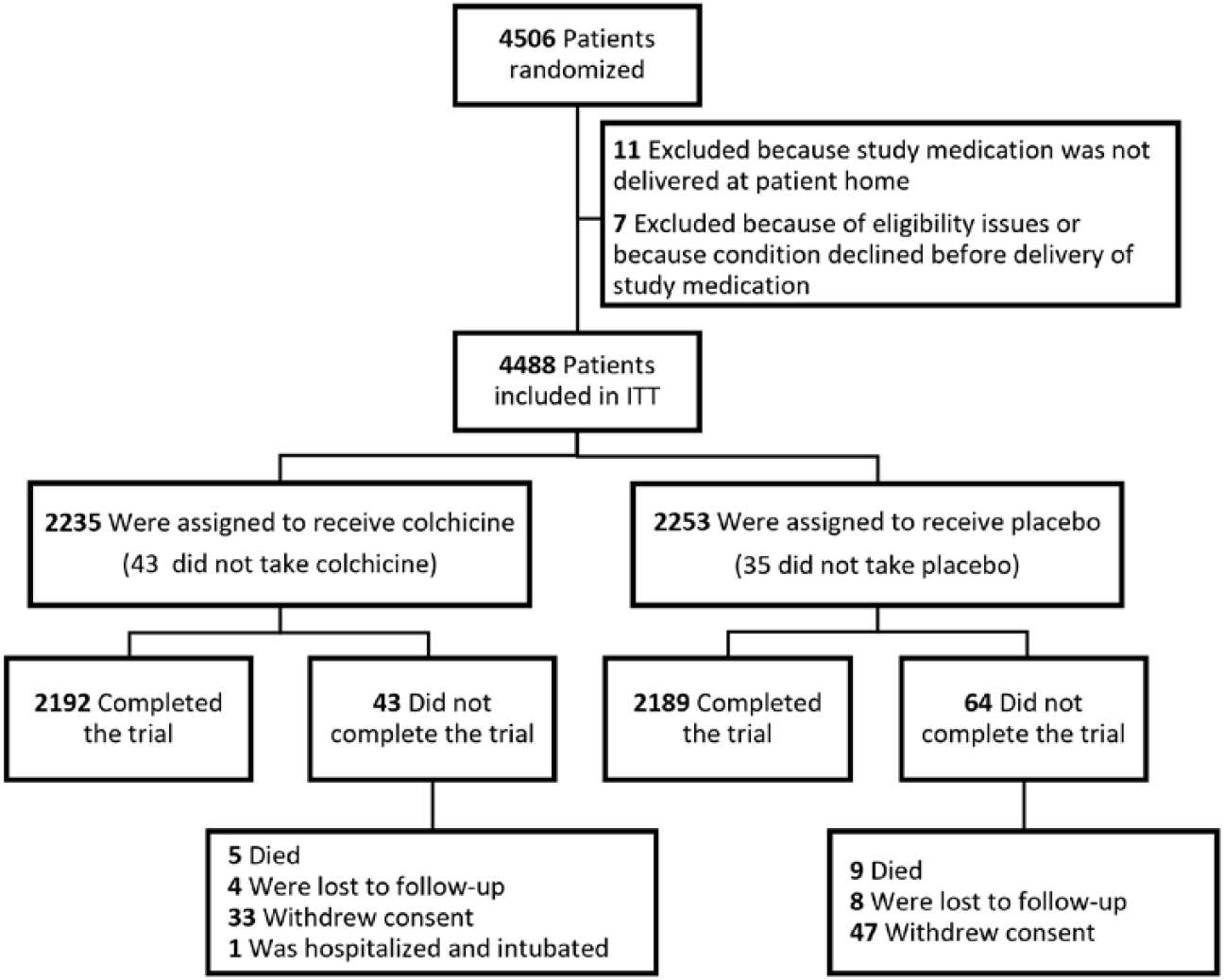
Consort Diagram of the Flow of Patients During the Trial.

The baseline characteristics of patients are shown in Table 1. Patients were enrolled a mean of 5.3 days after the onset of COVID-19 symptoms. The mean age of participants was 54.7 years, 53.9% of the patients were women, mean body-mass index was 30.0 kg/m^2^, and 19.9% had diabetes. The mean treatment duration with the trial medication was 26.2 days.

**Table 1.** Characteristics of the Trial Patients.

| **Characteristic** | **Colchicine (N=2235)** | **Placebo (N=2253)** |
| --- | --- | --- |
| Age - years | 54.4±9.7 | 54.9±9.9 |
| Female sex - no. (%) | 1238 (55.4%) | 1183 (52.5%) |
| Caucasian - no. (%) | 2086 (93.3%) | 2096 (93.2%) |
| Body-mass index (kg/m<sup>2</sup>) | 30.0±6.2 | 30.0±6.3 |
| Smoking - no. (%) | 217 (9.7%) | 212 (9.4%) |
| Hypertension - no. (%) | 781 (34.9%) | 848 (37.6%) |
| Diabetes - no. (%) | 444 (19.9%) | 450 (20.0%) |
| Respiratory disease - no. (%) | 583 (26.1%) | 605 (26.9%) |
| Prior MI - no. (%) | 65 (2.9%) | 72 (3.2%) |
| Prior heart failure - no. (%) | 24 (1.1%) | 18 (0.8%) |

MI denotes myocardial infarction.

### CLINICAL EFFICACY ENDPOINTS

A primary endpoint event occurred in 4.7% of the patients in the colchicine group, as compared with 5.8% of the patients in the placebo group (odds ratio, 0.79; 95.1% confidence interval [CI], 0.61 to 1.03; P=0.08). Table 2 shows the event rates and odds ratios for the components of the primary endpoint, which included death (odds ratio, 0.56; 95% CI, 0.19 to 1.67) and hospitalization due to COVID-19 (odds ratio, 0.79; 95% CI, 0.60 to 1.03), as well as for the secondary efficacy endpoint of the need for mechanical ventilation (odds ratio, 0.53; 95% CI, 0.25 to 1.09).

**Table 2.** Rates and Odds Ratios for Major Clinical Outcomes.

| **Clinical Outcome** | **Colchicine** | **Placebo** | **Odds Ratio**<br>**(95% CI)** | **P Value** |
| --- | --- | --- | --- | --- |
| <u>ITT population</u> | N=2235 | N=2253 |  |  |
| Primary composite endpoint - no. (%) | 104 (4.7%) | 131 (5.8%) | 0.79 (0.61-1.03) | 0.08 |
| Components of primary endpoint: |  |  |  |  |
| Death - no. (%) | 5 (0.2%) | 9 (0.4%) | 0.56 (0.19-1.67) |  |
| Hospitalization for COVID-19 no. (%) | 101 (4.5%) | 128 (5.7%) | 0.79 (0.60-1.03) |  |
| Secondary endpoint: |  |  |  |  |
| Mechanical ventilation - no. (%) | 11 (0.5%) | 21 (0.9%) | 0.53 (0.25-1.09) |  |
| <u>Patients with PCR-proven COVID-19</u> | N=2075 | N=2084 |  |  |
| Primary composite endpoint – no. (%) | 96 (4.6%) | 126 (6.0%) | 0.75 (0.57-0.99) | 0.04 |
| Components of primary endpoint: |  |  |  |  |
| Death – no. (%) | 5 (0.2%) | 9 (0.4%) | 0.56 (0.19-1.66) |  |
| Hospitalization for COVID-19 no. (%) | 93 (4.5%) | 123 (5.9%) | 0.75 (0.57-0.99) |  |
| Secondary endpoint: |  |  |  |  |
| Mechanical ventilation – no. (%) | 10 (0.5%) | 20 (1.0%) | 0.50 (0.23-1.07) |  |

ITT denotes Intention-to-Treat.

In the pre-specified analysis of the 4159 patients with COVID-19 confirmed by a polymerase chain reaction test, the rates of the primary endpoint were 4.6% and 6.0% in the colchicine and placebo groups, respectively (odds ratio, 0.75; 95% CI, 0.57 to 0.99; P=0.04). Among these patients with confirmed COVID-19, the odds ratios were 0.75 (95% CI, 0.57 to 0.99) for hospitalization due to the infection and 0.56 (95% CI, 0.19 to 1.66) for death. The secondary efficacy endpoint of the need for mechanical ventilation occurred in 0.5% of the patients in the colchicine group, as compared with 1.0% of the patients in the placebo group (odds ratio, 0.50; 95% CI, 0.23 to 1.07).

Efficacy results in prespecified subgroups are shown in Table 3. Among the patients with diabetes, the primary endpoint occurred in 6.1% of those in the colchicine group and 9.6% in the placebo group (odds ratio, 0.61; 95% CI, 0.37 to 1.01).

**Table 3.** Primary Efficacy Composite Endpoint in Prespecified Subgroups.

| **Subgroup** | **Colchicine** | **Placebo** | **Odds ratio**<br>**(95% CI)** |
| --- | --- | --- | --- |
| *no. of patients with event/total no. of patients (%)* |  |  |  |
| History of diabetes |  |  |  |
| Yes | 27/444 (6.1%) | 43/450 (9.6%) | 0.61 (0.37-1.01) |
| No | 77/1791 (4.3%) | 88/1803 (4.9%) | 0.88 (0.64-1.20) |
| History of hypertension |  |  |  |
| Yes | 48/781 (6.1%) | 64/848 (7.5%) | 0.80 (0.54-1.18) |
| No | 56/1454 (3.9%) | 67/1405 (4.8%) | 0.80 (0.56-1.15) |
| Smoking |  |  |  |
| Non-smoker | 59/1279 (4.6%) | 71/1270 (5.6%) | 0.82 (0.57-1.16) |
| Previous smoker | 38/738 (5.1%) | 56/770 (7.3%) | 0.69 (0.45-1.06) |
| Active smoker | 7/217 (3.2%) | 4/212 (1.9%) | 1.73 (0.50-6.01) |
| Age |  |  |  |
| ≥ 70 years | 18/190 (9.5%) | 27/213 (12.7%) | 0.72 (0.38-1.36) |
| < 70 years | 86/2045 (4.2%) | 104/2040 (5.1%) | 0.82 (0.61-1.09) |
| Sex |  |  |  |
| Men | 58/997 (5.8%) | 90/1070 (8.4%) | 0.67 (0.48-0.95) |
| Women | 46/1238 (3.7%) | 41/1183 (3.5%) | 1.07 (0.70-1.65) |
| Body-mass index |  |  |  |
| ≥ 30 kg/m<sup>2</sup> | 53/1012 (5.2%) | 70/1040 (6.7%) | 0.77 (0.53-1.11) |
| < 30 kg/m<sup>2</sup> | 50/1216 (4.1%) | 61/1205 (5.1%) | 0.80 (0.55-1.18) |
| Respiratory disease |  |  |  |
| Yes | 35/583 (6.0%) | 48/605 (7.9%) | 0.74 (0.47-1.16) |
| No | 69/1652 (4.2%) | 83/1647 (5.0%) | 0.82 (0.59-1.14) |
| Cardiovascular disease |  |  |  |
| Yes | 6/119 (5.0%) | 11/122 (9.0%) | 0.54 (0.19-1.50) |
| No | 98/2116 (4.6%) | 120/2131 (5.6%) | 0.81 (0.62-1.07) |
| Use of ACEi/ARB |  |  |  |
| Yes | 37/602 (6.1%) | 53/676 (7.8%) | 0.77 (0.50-1.19) |
| No | 67/1633 (4.1%) | 78/1577 (4.9%) | 0.82 (0.59-1.15) |

ACEi denotes angiotensin-converting enzyme inhibitor, and ARB angiotensin-receptor blocker.

### SAFETY AND ADVERSE EVENTS

The rates of serious adverse events were 4.9% in the colchicine group and 6.3% in the placebo group (P=0.05), and pneumonia occurred in 2.9% and 4.1% of the patients in the two groups (P=0.02). Pulmonary embolism was diagnosed in 0.5% of the patients in the colchicine group and 0.1% of those in the placebo group (0.01). The rates of adverse events that were considered related to trial medication were 24.2% and 15.5% (Table 4). At least one treatment-emergent gastro-intestinal adverse event occurred in 23.9% of the patients in the colchicine group, as compared with 14.8% of the patients in the placebo group. Diarrhea was reported in 13.7% and 7.3% of patients in the two trial groups (P<0.0001).

**Table 4.** Proportions of Patients with Adverse Events in the Safety Population†.

| **Adverse event** | **Colchicine (N=2195)** | **Placebo (N=2217)** | **P Value\*** |
| --- | --- | --- | --- |
| Any SAE - no. (%) | 108 (4.9%) | 139 (6.3%) | 0.05 |
| Pneumonia SAE - no. (%) | 63 (2.9%) | 92 (4.1%) | 0.02 |
| Pulmonary embolism - no. (%) | 11 (0.5%) | 2 (0.1%) | 0.01 |
| Any related AE - no. (%) | 532 (24.2%) | 344 (15.5%) | <0.0001 |
| Gastro-intestinal AE - no. (%) | 524 (23.9%) | 328 (14.8%) | <0.0001 |
| Gastro-intestinal SAE – no. (%) | 6 (0.3%) | 3 (0.1%) | 0.31 |
| Diarrhea AE - no. (%) | 300 (13.7%) | 161 (7.3%) | <0.0001 |
| Nausea AE - no. (%) | 43 (2.0%) | 47 (2.1%) | 0.71 |
| GI haemorrhage AE - no. (%) | 1 (0.0%) | 0 (0%) | 0.33 |
| Rash AE – no. (%) | 4 (0.2%) | 13 (0.6%) | 0.03 |

AE denotes adverse event; GI, gastro-intestinal; and SAE, serious adverse event.
†The safety population refers to the patients who took at least one dose of trial medication.

## DISCUSSION

In COLCORONA, the risk of the primary composite efficacy endpoint of death or hospitalization due to COVID-19 infection in the 30 days following randomization, was lower among the patients who were randomly assigned to receive colchicine than among those who received placebo. Because of the shortage of reagents for polymerase chain reaction tests and the restriction in the use of such testing early in the pandemic, diagnosis of probable COVID-19 through an epidemiological link or compatible symptoms was initially allowed in the study. When the 93% of patients who had a formal diagnosis of COVID-19 are considered, the benefit of colchicine on the primary efficacy endpoint was more marked (25%) and statistically significant. Treatment with colchicine was associated with concordant effects on hospitalizations, use of mechanical ventilation and deaths in patients with a diagnosis of COVID-19 confirmed by polymerase chain reaction testing.

The effect of colchicine on the primary endpoint was consistent across subgroups of patients based on various clinical characteristics. Although the benefits of colchicine appeared to be more marked in patients with diabetes and men, there was no significant heterogeneity in the results. Because the event rates were higher in patients with these characteristics, the effect of colchicine might have been more readily detectable. Diabetes is a pro-inflammatory state, which might explain the greater risk of complications of COVID-19 in patients afflicted by that disease. Despite the link between weight, insulin resistance and type 2 diabetes, the effects of colchicine did not differ whether the body-mass index was above or below 30 kg per square meter. In contrast, there is no readily obvious basis for a sex-related difference in responses to colchicine. Of note, the concomitant use of an inhibitor of the renin-angiotensin system did not appear to modify the clinical response to colchicine.

The most common adverse events observed were gastro-intestinal. Diarrhea was reported by 13.7% and 7.3% of patients in the colchicine and placebo groups, respectively. The number of patients with any serious adverse event was smaller in the colchicine group compared to placebo (4.9% versus 6.3%). Pneumonia was reported less frequently in patients of the colchicine group (2.9%) than those of the placebo group (4.1%). Colchicine has previously been shown to reduce acute lung injury in an experimental model of acute respiratory distress syndrome.^14^ The risk of viral inflammatory pneumonitis might therefore be lowered by colchicine in patients with COVID-19. In contrast, there was no evidence of an increased risk of bacterial pneumonia in COLCORONA. The number of reported cases of pulmonary embolism was higher in patients of the colchicine group compared to placebo (11 versus 2). Whether this represents a real phenomenon or simply the play of chance is not known. Colchicine has previously been shown in murine models to lower the release of alpha-defensin associated with large thrombus burdens and in clinical studies to reduce the aggregation between neutrophils and platelets.^16-18^ Nevertheless, the numbers of hospitalizations, use of mechanical ventilation and deaths were lower in the colchicine group than in the placebo group. Although uncommon, the number of reports of cutaneous rash was lower with colchicine than in the placebo group (4 versus 13).

Our trial has certain limitations. The study was stopped when 75% of the planned patients were recruited and had completed the 30-day follow-up. In addition to the logistical issues faced in the current challenging context, the perceived need to disseminate the study results rapidly in view of the current state of the pandemic largely contributed to our decision. The duration of follow-up was relatively short at approximately 30 days. The evolution of persistent COVID-19 symptoms and the effects of longer-term treatment with colchicine were not evaluated. The benefit of a shorter course of colchicine therapy for less than 30 days is also not entirely known, although the results of a small open-label study showed benefits of treatment administered for up to 3 weeks.^19^ Finally, our results apply to patients who have a proven diagnosis of COVID-19, are at risk of clinical complications and are not hospitalized at the time of treatment initiation.

In conclusion, among non-hospitalized patients with confirmed COVID-19, colchicine led to a lower rate of the composite of death or hospitalization than placebo.

## Supporting information

COI forms

## Data Availability

Data will be made available through vivli.org.

http://www.vivli.org

