## Supplementary material for "Efficacy of Colchicine in Non-Hospitalized Patients with COVID-19": COI forms: coi_disclosure_de_Luz.pdf

ICMJE INTERNATIONAL COMMITTEE of MEDICAL JOURNAL EDITORS

### ICMJE Form for Disclosure of Potential Conflicts of Interest

#### Section 1. Identifying Information

1. Given Name (First Name)
PROTASIO

2. Surname (Last Name)
da Luz

3. Date
25-January-2021

4. Are you the corresponding author? ☐ Yes ☑ No

Corresponding Author's Name
Jean-Claude Tardif

5. Manuscript Title
Efficacy of Colchicine in Non-Hospitalized Patients with COVID-19

6. Manuscript Identifying Number (if you know it)
MEDRXIV/2021/250494

#### Section 2. The Work Under Consideration for Publication

Did you or your institution **at any time** receive payment or services from a third party (government, commercial, private foundation, etc.) for any aspect of the submitted work (including but not limited to grants, data monitoring board, study design, manuscript preparation, statistical analysis, etc.)?

Are there any relevant conflicts of interest? ☐ Yes ☑ No

#### Section 3. Relevant financial activities outside the submitted work.

Place a check in the appropriate boxes in the table to indicate whether you have financial relationships (regardless of amount of compensation) with entities as described in the instructions. Use one line for each entity; add as many lines as you need by clicking the "Add +" box. You should report relationships that were **present during the 36 months prior to publication**.

Are there any relevant conflicts of interest? ☐ Yes ☑ No

#### Section 4. Intellectual Property -- Patents & Copyrights

Do you have any patents, whether planned, pending or issued, broadly relevant to the work? ☐ Yes ☑ No

### ICMJE Form for Disclosure of Potential Conflicts of Interest

#### Instructions

**The purpose of this form is to provide readers of your manuscript with information about your other interests that could influence how they receive and understand your work. The form is designed to be completed electronically and stored electronically. It contains programming that allows appropriate data display. Each author should submit a separate form and is responsible for the accuracy and completeness of the submitted information. The form is in six parts.**

##### 1. Identifying information.

##### 2. The work under consideration for publication.

This section asks for information about the work that you have submitted for publication. The time frame for this reporting is that of the work itself, from the initial conception and planning to the present. The requested information is about resources that you received, either directly or indirectly (via your institution), to enable you to complete the work. Checking "No" means that you did the work without receiving any financial support from any third party -- that is, the work was supported by funds from the same institution that pays your salary and that institution did not receive third-party funds with which to pay you. If you or your institution received funds from a third party to support the work, such as a government granting agency, charitable foundation or commercial sponsor, check "Yes".

##### 3. Relevant financial activities outside the submitted work.

This section asks about your financial relationships with entities in the bio-medical arena that could be perceived to influence, or that give the appearance of potentially influencing, what you wrote in the submitted work. You should disclose interactions with ANY entity that could be considered broadly relevant to the work. For example, if your article is about testing an epidermal growth factor receptor (EGFR) antagonist in lung cancer, you should report all associations with entities pursuing diagnostic or therapeutic strategies in cancer in general, not just in the area of EGFR or lung cancer.

Report all sources of revenue paid (or promised to be paid) directly to you or your institution on your behalf over the 36 months prior to submission of the work. This should include all monies from sources with relevance to the submitted work, not just monies from the entity that sponsored the research. Please note that your interactions with the work's sponsor that are outside the submitted work should also be listed here. If there is any question, it is usually better to disclose a relationship than not to do so.

For grants you have received for work outside the submitted work, you should disclose support ONLY from entities that could be perceived to be affected financially by the published work, such as drug companies, or foundations supported by entities that could be perceived to have a financial stake in the outcome. Public funding sources, such as government agencies, charitable foundations or academic institutions, need not be disclosed. For example, if a government agency sponsored a study in which you have been involved and drugs were provided by a pharmaceutical company, you need only list the pharmaceutical company.

##### 4. Intellectual Property.

This section asks about patents and copyrights, whether pending, issued, licensed and/or receiving royalties.

##### 5. Relationships not covered above.

Use this section to report other relationships or activities that readers could perceive to have influenced, or that give the appearance of potentially influencing, what you wrote in the submitted work.

##### Definitions.

**Entity:** government agency, foundation, commercial sponsor, academic institution, etc.
**Grant:** A grant from an entity, generally [but not always] paid to your organization
**Personal Fees:** Monies paid to you for services rendered, generally honoraria, royalties, or fees for consulting , lectures, speakers bureaus, expert testimony, employment, or other affiliations
**Non-Financial Support:** Examples include drugs/equipment supplied by the entity, travel paid by the entity, writing assistance, administrative support, etc.

**Other:** Anything not covered under the previous three boxes
**Pending:** The patent has been filed but not issued
**Issued:** The patent has been issued by the agency
**Licensed:** The patent has been licensed to an entity, whether earning royalties or not
**Royalties:** Funds are coming in to you or your institution due to your patent

ICMJE INTERNATIONAL COMMITTEE *of* MEDICAL JOURNAL EDITORS

### ICMJE Form for Disclosure of Potential Conflicts of Interest

#### Section 5. Relationships not covered above

Are there other relationships or activities that readers could perceive to have influenced, or that give the appearance of potentially influencing, what you wrote in the submitted work?

☐ Yes, the following relationships/conditions/circumstances are present (explain below):

☑ No other relationships/conditions/circumstances that present a potential conflict of interest

At the time of manuscript acceptance, journals will ask authors to confirm and, if necessary, update their disclosure statements. On occasion, journals may ask authors to disclose further information about reported relationships.

#### Section 6. Disclosure Statement

Based on the above disclosures, this form will automatically generate a disclosure statement, which will appear in the box below.

#### Evaluation and Feedback

Please visit http://www.icmje.org/cgi-bin/feedback to provide feedback on your experience with completing this form.
