## Supplementary material for "Efficacy of Colchicine in Non-Hospitalized Patients with COVID-19": COI forms: coi_disclosure_Dubois.pdf

To view the full contents of this document, you need a later version of the PDF viewer. You can upgrade to the latest version of Adobe Reader from www.adobe.com/products/acrobat/readstep2.html

For further support, go to www.adobe.com/support/products/acrreader.html
